# CerViX-Net: A Multi-Branch Fusion of Vision Transformer and Convolutional Neural Networks for Cervical Cancer Detection using Cytology Images

**DOI:** 10.64898/2026.06.24.26356425

**Authors:** Sagnik De

## Abstract

Cervical cancer represents a pressing global health challenge, emphasizing the critical need for accurate and timely diagnostic methods to facilitate effective treatment and improve survival rates. In response to this challenge, the study presents CerViX-Net, an innovative classification framework designed to advance cervical cancer detection through enhanced computational efficiency and diagnostic accuracy. The development of CerViX-Net is motivated by the limitations of traditional diagnostic models, particularly in handling the computational and memory demands of large-scale data, while ensuring precise feature extraction and classification. CerViX-Net employs a hybrid deep learning architecture that combines the capabilities of ResNet50, EfficientNet-B0, and a Modified Vision Transformer (ViT) module. The ResNet50 branch extracts hierarchical features through stacked convolutional and identity blocks. In another path, the modified ViT module transforms image patches via linear projection, augments them with positional and class embeddings, and processes them using Parallel Transformer Encoder layers to model contextual relationships. Concurrently, EfficientNet-B0 utilizes MBConv blocks to extract multi-scale representations. The feature outputs from all three branches are integrated and passed through a classification head consisting of dropout layers and dense layers to ensure robust and accurate predictions. The proposed framework is rigorously evaluated on the Mendeley LBC dataset, achieving exceptional performance metrics with an accuracy of 99.69%, precision of 99.28%, recall of 99.48%, and an F1-score of 99.52%. The robustness of CerViX-Net is further validated on the SIPaKMeD and Herlev Pap Smear datasets, where it demonstrates comparable excellence, underscoring its efficacy and adaptability across diverse cytology datasets. Statistical validation using Friedman’s test further reinforces its superiority over competing methods.

## I. Introduction

Cervical cancer, ranking as the fourth most common cancer among females globally, poses a significant health challenge, with nearly 604,000 new cases and 342,000 deaths recorded in 2020 alone [1]. The burden of this disease disproportionately affects low- and middle-income countries, where about 90% of cases and deaths occur. The cervix, a thin tissue layer covering a woman’s reproductive tract, can undergo malignant transformation over several years, progressing from a precancerous lesion stage to invasive cancer if left unchecked [2]. However, timely identification at the pre-cancerous phase offers opportunities for effective treatment and prevention of further progression. The Pap smear test is the most widely used screening method for detecting cervical cancer. This cytology examination involves the manual review of Pap smear slides, where cytopathologists examine numerous micro-images to identify abnormalities. This process is labor-intensive and prone to human error, as subtle irregularities can be missed. The complexity of Pap smear images [5], where small precancerous lesions may be hidden by overlapping cells or mucus, adds to the challenge, making the procedure highly error-prone [3].

Intelligent systems and medical image processing technologies provide cost-effective and time-efficient alternatives to traditional screening methods such as Pap smear, colposcopy, and cervicography. Advancements in computer-aided diagnostic (CAD) techniques [4] offer promising solutions to these challenges by automating cancer detection and improving accuracy. These systems assist healthcare professionals by enhancing image analysis, identifying subtle abnormalities, and reducing diagnosis variability.

In early 2000, cervical lesion detection based on machine learning (ML) techniques started to be extensively researched. Recent developments in CAD systems aim to enhance the effectiveness of cervical cancer screening by improving image quality, segmenting regions of interest, and identifying abnormal patterns [9]. These tools are particularly beneficial in colposcopy, where they can highlight pathological areas such as acetowhite regions, abnormal vascularization, mosaic patterns, and punctation [10]. By providing valuable diagnostic support, CAD systems help clinicians make more accurate decisions, potentially leading to better patient outcomes and reduced mortality rates. Traditional ML requires a feature engineering process and needs to extract pre-selected features that are used for model learning.

Traditional diagnostic models often struggle with computational complexity, memory overhead, and limitations in feature extraction, especially when applied to large-scale datasets. These shortcomings highlight the necessity for innovative approaches that balance computational efficiency with robust diagnostic accuracy, ensuring practical applicability in clinical and resource-constrained settings. Motivated by these challenges, this study aims to develop CerViX-Net, a novel classification framework designed to enhance cervical cancer diagnosis. The objectives are threefold: (i) to address the computational and memory demands of large-scale data analysis, to refine feature extraction processes for more precise classification, and (iii) to ensure the adaptability and robustness of the framework across diverse cytological datasets.

The proposed CerViX-Net framework is structured around a multi-branch deep learning architecture that integrates three complementary modules to maximize feature diversity and classification accuracy. The first component leverages the ResNet50 backbone, which extracts layered spatial features through sequential convolutional and identity blocks. In parallel, a Modified Vision Transformer (ViT) module segments input images into patches, applies linear projections along with positional and class embeddings, and routes the embedded tokens through Parallel Transformer Encoder layers to capture intricate contextual relationships. The third branch utilizes EfficientNet-B0, known for its efficient scaling and depthwise separable convolutions, to extract multi-scale semantic features via MBConv blocks. Outputs from all three branches are concatenated into a unified representation, which is then processed through a fully connected classification head composed of dropout layers and dense layers. This integrated design enables CerViX-Net to jointly leverage deep spatial hierarchies, attention-based context modeling, and efficient multiscale feature extraction for robust classification of cervical cytology images.

The key contributions of this study include:

1. Design of CerViX-Net, a three-branch architecture that integrates a ResNet50 module for deep feature extraction, a Modified ViT module with parallel transformer encoders for global context learning, and an Efficient-NetB0 module for efficient multi-scale representation.
2. Incorporation of parallel transformer encoders within the ViT module, enabling more efficient global feature extraction by addressing the limitations of traditional sequential transformer encoders.
3. Simultaneous feature learning from three complementary pathways—ResNet for deep residual learning, EfficientNet for lightweight and scalable representation, and the ViT module for capturing global dependencies—resulting in a richer and more discriminative feature space.
4. Evaluation of the proposed framework on three publicly available datasets: the SIPaKMeD Pap Smear dataset, the Herlev Pap Smear dataset, and the Mendeley Liquid-Based Cytology dataset, achieving classification accuracies of 99.68%, 98.36%, and 99.72%, respectively.

The flow of this article is as follows. Section II details the literature review, Section III presents an in-depth explanation of the methodology and architecture used in the CerViX-Net model, Section IV depicts the experiments and empirical results, while Section V provides the discussion regarding the proposed work. Finally, Section VI concludes the study.

## II. Literature Survey

Several studies have been conducted on the identification of Cervical Cancer cells. Plissiti et al. [2] delineated cervical cancer cell classification for the first time using SiPaKMeD dataset through MLP, SVM and CNN, achieving an accuracy of approximately 95.35% using CNN model. Win et al. [5] proposed a computer-assisted screening technique for Pap smear images, employing digital image processing and achieving an accuracy of 94.09% by leveraging texture, shape, and color features. Tripathi et al. [3] employed several CNN models, including ResNet 50, ResNet 152, VGG16, and VGG19, to classify cervical cancer cells. Among these models, ResNet 152 obtained the highest classification accuracy at 94.89%. Also, three CNN architectures—Inception v3, Xception, and DenseNet-169—that were pre-trained on the ImageNet dataset were combined to create an ensemble-based classification model by Manna et al. [4] Classifying Pap stained single cell and whole-slide images is done using this methodology. By applying two non-linear functions to the decision scores from the basis learners, the ensemble applies a fuzzy rank-based fusion of classifiers. The model performed well on the SIPaKMeD Pap Smear dataset, with 98.55% classification accuracy and 98.52% sensitivity in the 2-class setting and 95.43% accuracy and 98.52% sensitivity for 5-class setting. Basak et al. [12] introduced a framework utilizing feature selection and deep learning (DL) with evolutionary optimization for cytology image categorization, achieving an accuracy of 97.87%. Furthermore, Alquran et al.[10], have proposed an automated system for cervical cancer utilizing a novel DL architecture to obtain and fuse significant features. Achieving the highest accuracy of 99.1% using a SVM classifier, the proposed system selected 544 most significant features from Cervical Net and combined them with 544 from Shuffle Net, demonstrating superior performance for classifying five categories. Lastly, Mulmule et al. [13] put forward a method employing adaptive fuzzy k-means clustering to segment cells from pathological Pap smear images, extracting 40 features based on various cellular characteristics. In their work, a supervised classification approach utilizing a Multilayer Perceptron (MLP) and Support Vector Machine (SVM) classifiers demonstrated comparable performance to existing techniques. Trained and tested on a database with 280 Pap smear images, the MLP classifier, employing tanh, outperformed the SVM classifier with a accuracy of 97.14%, sensitivity of 98%, specificity of 95%, and positive predictive rate of 98%.

Also exploring works based on other datasets, Bora et al. [20] achieved 96.5% accuracy using an Ensemble classifier on the Herlev Dataset. Chankong et al. [14] demonstrated that among various classifiers for binary and multi-class classification, the ANN achieved the highest accuracy of 93.78% on same dataset. Similarly, Zhang et al. [15] utilized a deep convolutional neural network, which eliminated the need for cell segmentation and yielded a high accuracy of 98.3%. Finally, Marinakis et al. [16] applied a genetic algorithm combined with a nearest neighbour classifier, achieving an impressive 98.14% accuracy for the 2-class problem and 96.95% for the 7-class problem in 10-fold cross-validation.

Liming Hu et al. [6] developed a DL-based visual evaluation algorithm to classify cervical cancer. A faster R-CNN-based approach was used in this paper and achieved an accuracy rate of 91%. Kumar et al. [7] proposed a fusion-based DL approach to classify cervical cancer. Several DL architectures such as AlexNet, ResNet-152, ResNet-101, and Inception V3 were used to extract the features from the images, and used several ML classifiers to classify the images. The SIPaKMeD dataset used in this work achieved an accuracy rate of 98.08%. Kalbhor et al. [8] highlighted deep learning’s impact through two strategies. ResNet-50 achieved 92.03% accuracy as a feature extractor paired with machine learning. Transfer learning with GoogleNet excelled, reaching 96.01% classification accuracy.

Furthermore, Singh et al. [17] investigated cervical cancer diagnosis using various machine learning algorithms on benchmark datasets. Hybrid approaches were applied to address segmentation issues, while tree classifiers optimized feature selection. Certain algorithms exceeded 100% in metrics like accuracy and F1 scores with higher training data proportions. Although logistic regression with L1 regularization reached 100% accuracy, it demanded more computational resources compared to alternatives achieving 99% accuracy with reduced CPU usage. Kumawat et al. [18] employed supervised learning techniques on the UCI cervical cancer dataset. Models were evaluated with and without feature selection methods, including Relief rank, wrapper approaches, and LASSO regression. XGBoost delivered 94.94% accuracy using all features, while feature selection occasionally yielded better results.

This study addresses several critical limitations identified in existing cervical cancer classification models. Table I summarises the related works in terms of method used, dataset, performances along with their limitations. Many current approaches achieve high accuracy but struggle with generalization across diverse image conditions and varying noise levels, limiting their robustness. Additionally, the heavy reliance on pre-trained models hinders adaptability to new datasets and domain-specific variations. Furthermore, most methods focus on a narrow subset of features, neglecting the potential of high-dimensional feature sets that could enhance model performance. Lastly, while hybrid machine learning approaches improve accuracy, they often incur high computational costs, necessitating more efficient, resource-conserving techniques for practical deployment. This study aims to bridge these gaps with an innovative approach.

**TABLE I.** Summarization of related works.

| Author | Preprocessing | Model Used | Dataset | Performance | Limitations |
| --- | --- | --- | --- | --- | --- |
| Plissiti et al. [2] | - | CNN with SVM and MLP | SIPaKMeD dataset | Acc = 94.44% (SVM) | Extraction of handcrafted features is time consuming |
| Win et al. [5] | Noise removal, Nuclei and Cytoplasm segmentation | Handcrafted feature extraction, Feature selection, and classification | SIPaKMeD dataset<br>Herlev dataset | Acc = 90.84%<br>Acc = 94.09% | Extraction and manual features and selection of appropriate features is challenging |
| Tripathi et al. [3] | - | Multiple CNN architecture | SIPaKMeD dataset | Acc = 94.89% (ResNet-152) | Pre-trained models were used only |
| Alquran et al. [10] | Augmentation | ResNet101+SVM | Herlev dataset | AUC = 97.3% | Limited dataset were used |
| Mulmule et al. [13] | adaptive fuzzy k means clustering | Handcrafted features with MLP and SVM classifier |  | Auc = 97.14% (MLP) | Extraction of proper set of features is complex |
| Liming Hu et al. [6] | Annotation | Faster RCNN | 2000 Captured image | AUC = 91% | Limited to the smaller number of cases |
| Mathivanan et al. [7] | - | Fusion of multiple DL architecture with RF, PCA, DT, NB | SIPaKMeD dataset | Acc = 98.08% | Combination of multiple architecture increases the model complexity |
| Kalbhor et al. [8] | Image resizing | Hybridization of DL architectures | Herlev dataset | Acc = 96.01% (GoogleNet) | Limited dataset were used |

## III. Materials and Methododology

The flow of this study begins with the selection of three cytology datasets—Mendeley LBC, SIPaKMeD, and Herlev Pap Smear—for their diverse imaging characteristics. (i) Image refinement techniques are applied to remove noise and enhance cellular feature clarity, ensuring high-quality inputs for analysis. (ii) The refined images are then processed through CerViX-Net, which integrates three distinct modules: a ResNet50-based convolutional branch, an EfficientNetB0 module for efficient multi-scale representation, and a redesigned Vision Transformer (ViT) module with Parallel Transformer Encoders for effective global feature extraction. (iii) The Parallel Transformer Encoder structure enables simultaneous multi-head attention computation across patch embeddings, improving efficiency and contextual understanding compared to traditional sequential transformers. (iv) Feature outputs from all three branches are flattened and passed through dense layers for classification. (v) This comprehensive workflow allows CerViX-Net to achieve high accuracy in classifying cervical cytology images into distinct diagnostic categories. The workflow of this study is illustrated in Fig. 1.

**Fig. 1.**
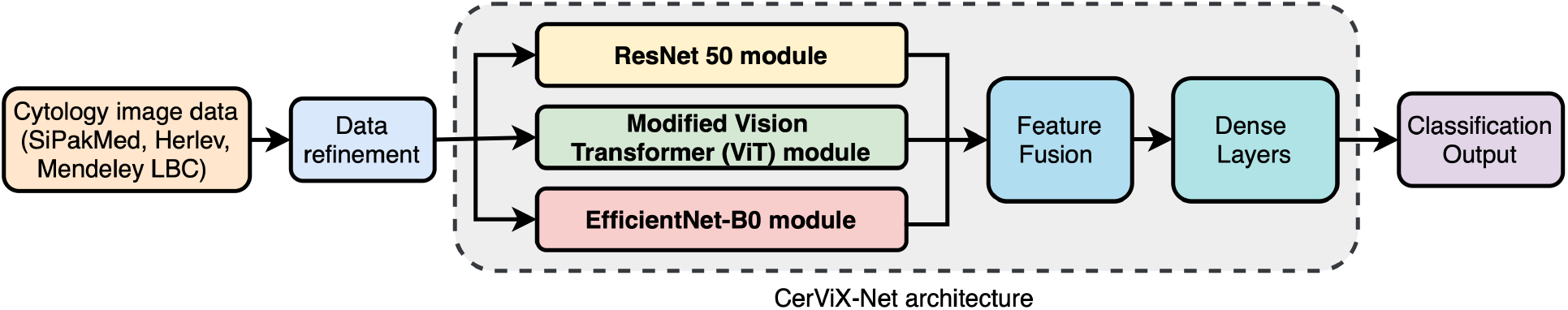
Schematic overview of the proposed workflow for Cervical Cancer detection.

### A. Dataset

#### 1) SiPakMed Dataset

This study employs the SiPakMed dataset [2], a renowned resource for cervical cell examination research, with Pap smear images being a primary method utilized. Publicly available datasets comprising cluster cervical cell images are numerous, and among them, the SIPaKMeD dataset stands out. It encompasses five distinct cell categories: superficial-intermediate, parabasal, koilocytotic, metaplastic, and dyskeratotic. Obtained from Pap smears captured via a CCD camera connected to a microscope, this dataset consists of 966 clusters of cell images, which have been snipped to yield 4049 individual cell images. Each image is meticulously labeled, assigning it to one of the five specified categories. While superficial-intermediate and parabasal cells represent typical cell types, koilocytotic and dyskeratotic cells indicate abnormalities without malignancy. Additionally, the last category comprises benign cells, specifically identified as metaplastic.

#### 2) Herlev Pap Smear Dataset

The Herlev Pap Smear Dataset [12] is a widely used collection for the classification of cervical cell images. It consists of 917 images, categorized into seven distinct classes based on cell type and health status. The classes include three Normal classes: Intermediate Squamous Epithelial, Columnar Epithelial, and Superficial Squamous Epithelial; and four Abnormal classes: Mild Squamous nonkeratinizing Dysplasia, Squamous cell carcinoma in-situ intermediate, Moderate Squamous non-keratinizing Dysplasia, and Severe Squamous non-keratinizing Dysplasia. This dataset is instrumental in developing and validating automated diagnostic systems for cervical cancer screening.

#### 3) Mendeley LBC Dataset

The Mendeley LBC (Liquid-Based Cytology) Dataset [12] is a comprehensive collection of cervical cytology images, containing 963 images categorized into four classes. The dataset is used to support research in cervical cancer diagnosis and screening. The classes include one Normal class: Negative for Intraepithelial Malignancy, and three Abnormal classes: Low grade Squamous Intraepithelial Lesion (LSIL), High grade Squamous Intraepithelial Lesion (HSIL), and Squamous Cell Carcinoma (SCC). The dataset facilitates the development of machine learning algorithms for the early detection of precancerous and cancerous lesions in cervical cells.

A sample from each class across all three datasets is presented in Fig. 2, while Table II provides an overview of the datasets.

**Fig. 2.**
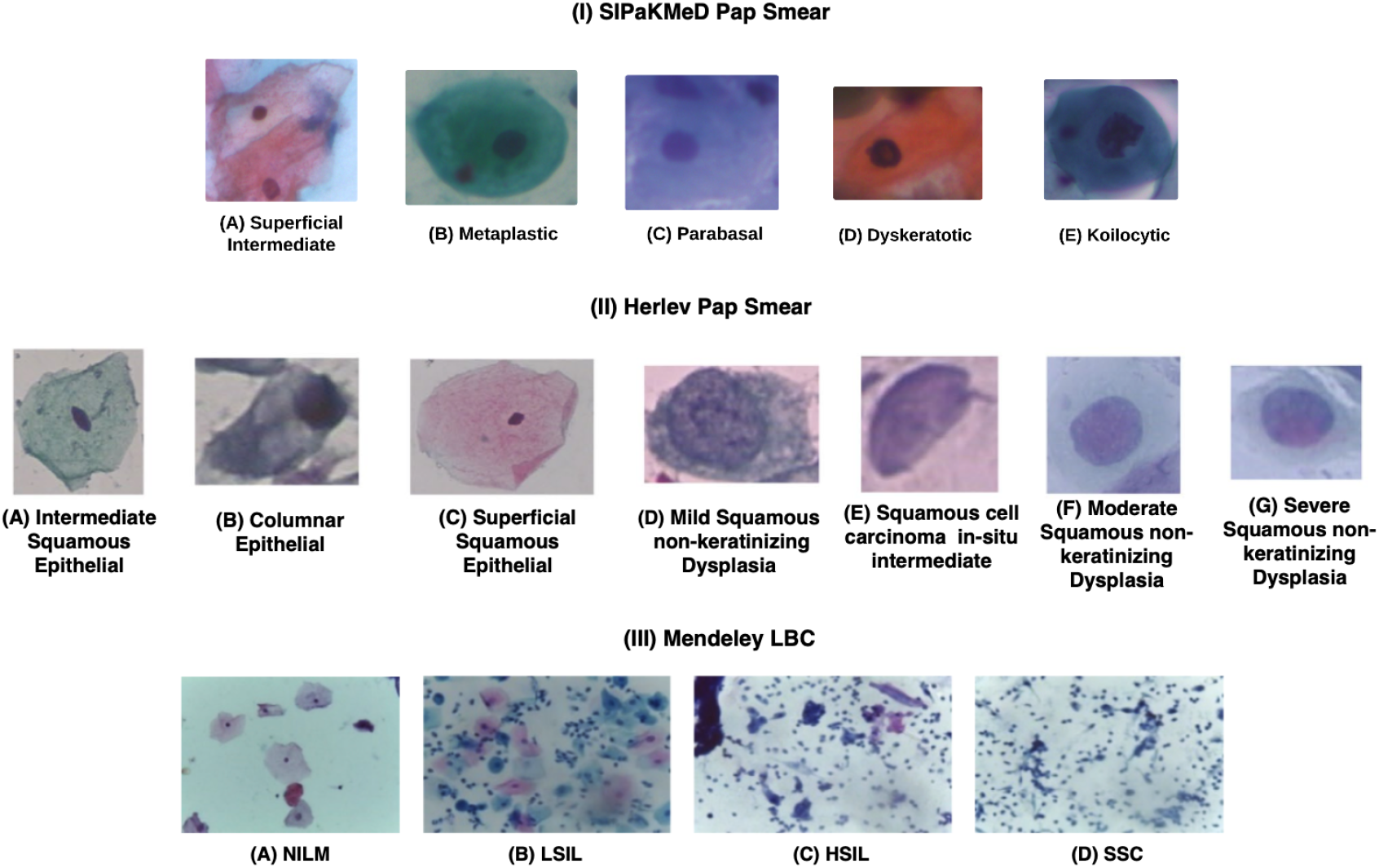
Representative images from datasets: (I) SIPaKMeD Pap Smear, (II) Herlev Pap Smear, and (III) Mendeley LBC.

**TABLE II.** Overview of the datasets considered in this study.

| Dataset | Class | Category | Cell Type | Number of Images |
| --- | --- | --- | --- | --- |
| SiPaKMeD Pap Smear (4049 images) | 1 | Normal | Superficial-Intermediate (SI) | 831 |
|  | 2 | Normal | Parabasal (PB) | 787 |
|  | 3 | Abnormal | Koilocytotic (KC) | 825 |
|  | 4 | Benign | Metaplastic (MT) | 793 |
|  | 5 | Abnormal | Dyskeratotic (DK) | 813 |
| Herlev Pap Smear (917 images) | 1 | Normal | Intermediate Squamous Epithelial (ISE) | 70 |
|  | 2 | Normal | Columnar Epithelial (CE) | 98 |
|  | 3 | Normal | Superficial Squamous Epithelial (SSE) | 74 |
|  | 4 | Abnormal | Mild Squamous non-keratinizing Dysplasia (MSND) | 182 |
|  | 5 | Abnormal | Squamous cell carcinoma in-situ intermediate (SCCI) | 150 |
|  | 6 | Abnormal | Moderate Squamous non-keratinizing Dysplasia (MoSND) | 146 |
|  | 7 | Abnormal | Severe Squamous non-keratinizing Dysplasia (SSND) | 197 |
| Mendeley LBC (963 images) | 1 | Normal | Negative for Intraepithelial Malignancy (NIM) | 613 |
|  | 2 | Abnormal | Low grade Squamous Intraepithelial Lesion (LSIL) | 163 |
|  | 3 | Abnormal | High grade Squamous Intraepithelial Lesion (HSIL) | 113 |
|  | 4 | Abnormal | Squamous Cell Carcinoma (SCC) | 74 |

### B. Image refinement

Medical images, such as Pap smear slides, often contain significant noise, which can affect the quality and accuracy of analysis. To address this challenge, the proposed model employs a series of preprocessing steps to reduce noise and enhance image quality. Initially, a median filter with a 3 × 3 kernel size is applied, effectively removing noise while preserving important edge details, which is more efficient than traditional convolution filters. Following noise reduction, histogram equalization [19] and normalization techniques are used to improve contrast by stretching the intensity histogram across a broader range. This contrast enhancement facilitates the extraction of more distinguishable features, which improves classification accuracy. These preprocessing steps ensure that the model works with clearer, more informative images, leading to superior performance.

### C. CerViX-Net architecture

#### 1) ResNet-50 module

The ResNet-50 architecture is a deep convolutional neural network consisting of 50 layers, structured using residual learning blocks. It begins with stage 1, which includes a 7 × 7 convolution layer with 64 filters and a stride of 2, followed by batch normalization, a ReLU activation function, and a 3 × 3 max pooling layer with a stride of 2. This reduces the input spatial dimensions and sets the stage for feature extraction. Stage 2 starts with a convolutional block that includes three convolutional layers: a 1 × 1 convolution with 64 filters, a 3 × 3 convolution with 64 filters, and a 1 × 1 convolution with 256 filters. This is followed by an identity block with the same layer configuration. Stage 3 increases the filter depth, starting with a convolutional block consisting of a 1 × 1 convolution with 128 filters, a 3 × 3 convolution with 128 filters, and a 1 × 1 convolution with 512 filters, followed by an identity block with identical dimensions. Stage 4 further increases the depth with a convolutional block using 256, 256, and 1024 filters in the 1 × 1, 3 × 3, and 1 × 1 convolutions respectively, followed by a matching identity block. Stage 5 has a 1 × 1, 3 × 3, and 1 × 1 convolutional block with 512, 512, and 2048 filters, followed by an identity block of the same shape. The architecture concludes with a global average pooling layer, which reduces the final feature map to a feature vector used for classification.

#### 2) Proposed modified ViT module

Given an input image *I* ∈ ℝ^*H×W ×Z*^, where *H* and *W* are the height and width of the image, and *Z* is the number of channels, the image is first divided into a grid of non-overlapping patches. Each patch *p* is of size *P* × *P* × *Z* [28]. The total number of patches is given by:

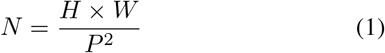

Each patch is then flattened into a vector and projected into a linear embedding space using a learnable matrix *E* [28]. The embedding process is mathematically represented as:

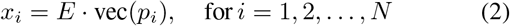

where vec(*p*_*i*_) denotes the flattened vector of the *i*-th patch, and *x*_*i*_ ∈ ℝ^*D*^ is the corresponding embedded vector.

After embedding the patches, a learnable class token *x*_class_ ∈ ℝ^*D*^ is appended to the sequence of embedded patches. This token will ultimately aggregate information from the entire sequence, acting as a global representation of the image.

Additionally, positional embeddings *E*_pos_ ∈ ℝ^(*N* +1)*×D*^ are added to the patch embeddings to retain the positional information, as the Transformer architecture is inherently permutation-invariant. The final input sequence to the Transformer encoder is:

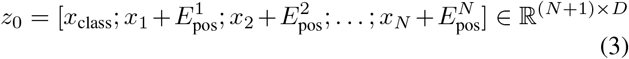

Multi-Head Attention (MHA) is a key component of the Vision Transformer, enabling the model to jointly attend to information from different representation subspaces. Given an input sequence *X* ∈ ℝ^*n×d*^, where *n* is the number of tokens and *d* is the embedding dimension, MHA projects *X* into queries *Q*, keys *K*, and values *V* using learned matrices:

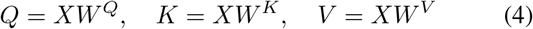

where, 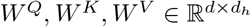 and 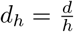 for *h* attention heads.

The scaled dot-product attention for each head is:

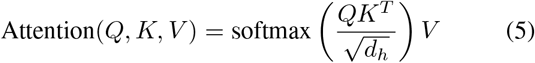

For *h* heads, the outputs are concatenated and linearly transformed:

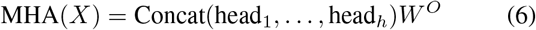

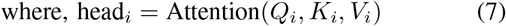

Here, *W*^*O*^ ∈ ℝ^*d×d*^ is the output projection matrix. This mechanism enables the model to capture complex relationships between patches across multiple representation subspaces simultaneously.

A standard transformer block processes its input through sequential operations of multi-head self-attention (MHA), layer normalization (LN), and a multi-layer perceptron (MLP), with residual connections at each stage. If *Z*_*m*_ denotes the output of the *m*-th transformer layer, then the transformation across layers can be expressed through the following steps:

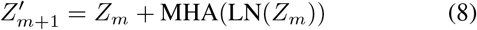

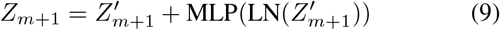

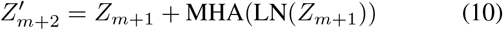

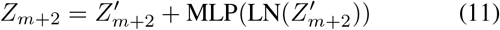

Here, *m* ∈ { 1, …, *N* } indexes the layers. To improve learning effectiveness and reduce redundancy across deep transformer stacks, the Parallel Transformer Encoder (PE) is introduced. Instead of stacking two sequential transformer blocks, PE combines their operations in parallel within a single layer.

In the parallel version, the input *Z*_*m*_ is processed through two MHA modules simultaneously after applying layer normalization, and their outputs are summed along with the residual connection:

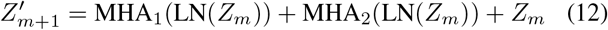

The resulting intermediate 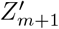 is then passed through two parallel MLP branches, also normalized, and their outputs are added along with a residual connection from 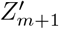.

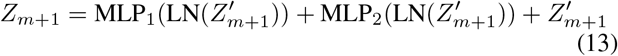

This architecture reduces the model’s depth while preserving representational power, aiming to enhance learning efficiency and mitigate diminishing effects of residual paths in deeper stacks. The proposed parallel transformer encoder based ViT architecture is illustrated as in Fig. 3.

**Fig. 3.**
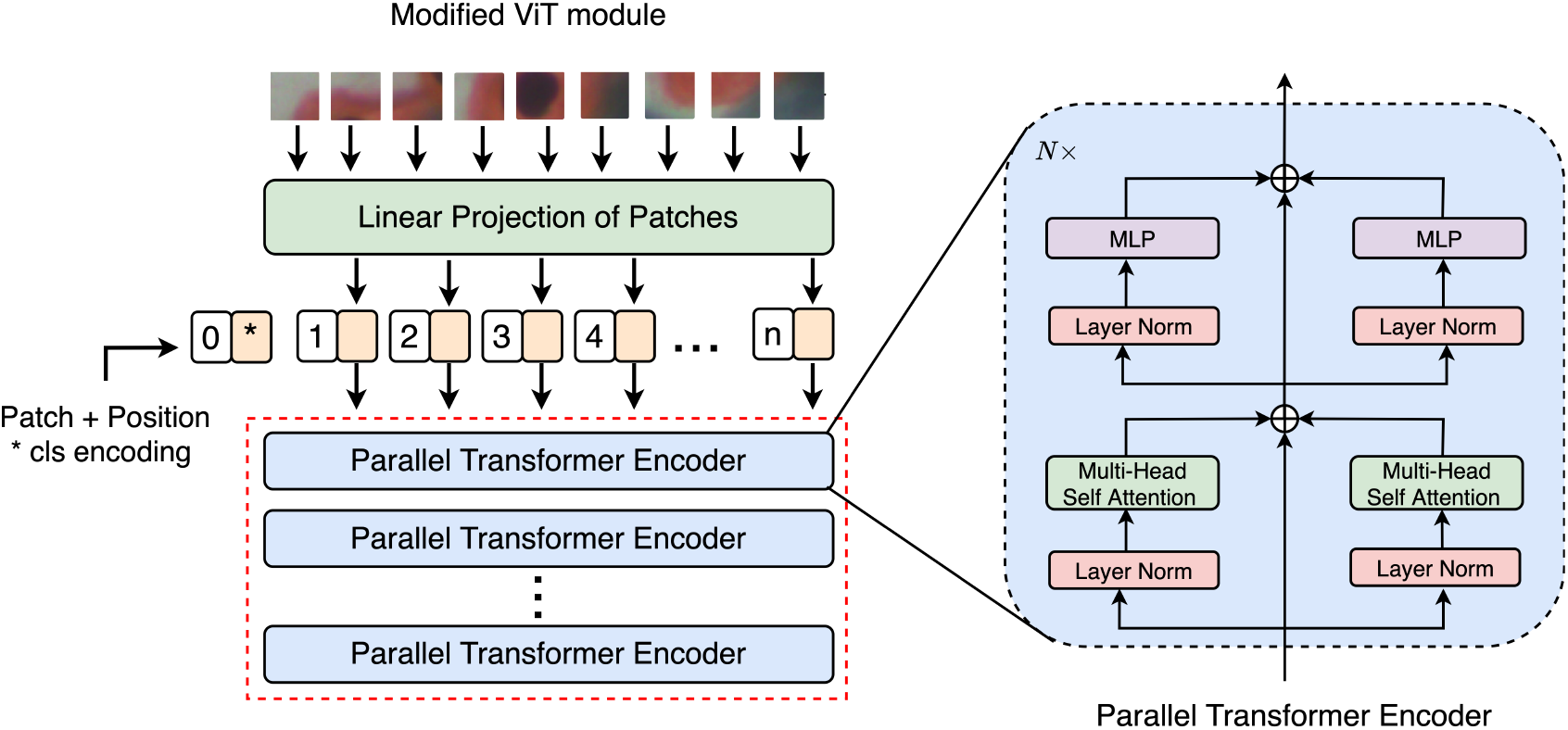
Visualization of the proposed parallel transformer encoder incorporated in the modified ViT module.

#### 3) EfficientNet-B0 module

The EfficientNet-B0 module shown above begins with a standard 3 × 3 convolutional layer that extracts low-level features from the input image. Following this, the network is composed of sequential blocks containing MBConv layers, which stand for Mobile Inverted Bottleneck Convolutions. These layers are denoted as MBConv*X, Y* × *Y*, where *X* indicates the expansion factor and *Y* × *Y* specifies the kernel size. Block 1 contains a single MBConv1, 3 × 3. Block 2 introduces two MBConv6, 3 × 3 layers, increasing the model’s representational capacity. In Block 3, the network uses two MBConv6, 5 × 5 layers, enabling it to capture broader spatial features. Block 4 adds three MBConv6, 3 × 3 layers to deepen the network. Block 6, which is the longest, includes five MBConv6, 5 × 5 layers, significantly enhancing the depth and complexity. Finally, Block 7 contains a single MBConv6, 3 × 3, preparing features for classification. Each MBConv layer consists of a pointwise expansion, a depthwise convolution, and a pointwise projection, often accompanied by squeeze-and-excitation mechanisms and residual connections. This hierarchical and efficient design allows EfficientNet-B0 to achieve high performance with reduced computational cost.

#### 4) Overall flow of CerViX-Net architecture

The outputs from the ResNet50, EfficientNetB0, and modified ViT branches are fused to form a unified feature vector. Let the respective outputs be *F*_Res_, *F*_Eff_, *F*_ViT_, which are concatenated as

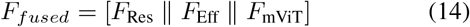

This combined feature representation captures local, hierarchical, and global contextual information. The fused vector *F* is passed through a classification head comprising two dense layers interleaved with dropout for regularization. Mathematically, the final prediction *ŷ* is computed as:

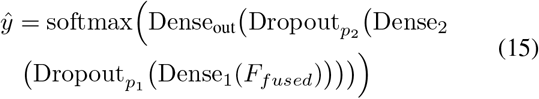

This design enables robust and accurate multi-class classification by minimizing overfitting while leveraging the complementary strengths of the three backbone networks. The final output layer adapts to the dataset’s class configuration, supporting classification into 5 (SIPaKMeD), 7 (Herlev), or 4 (Mendeley LBC) classes. The overall architecture of CerViX-Net is as depicted in Fig. 4.

**Fig. 4.**
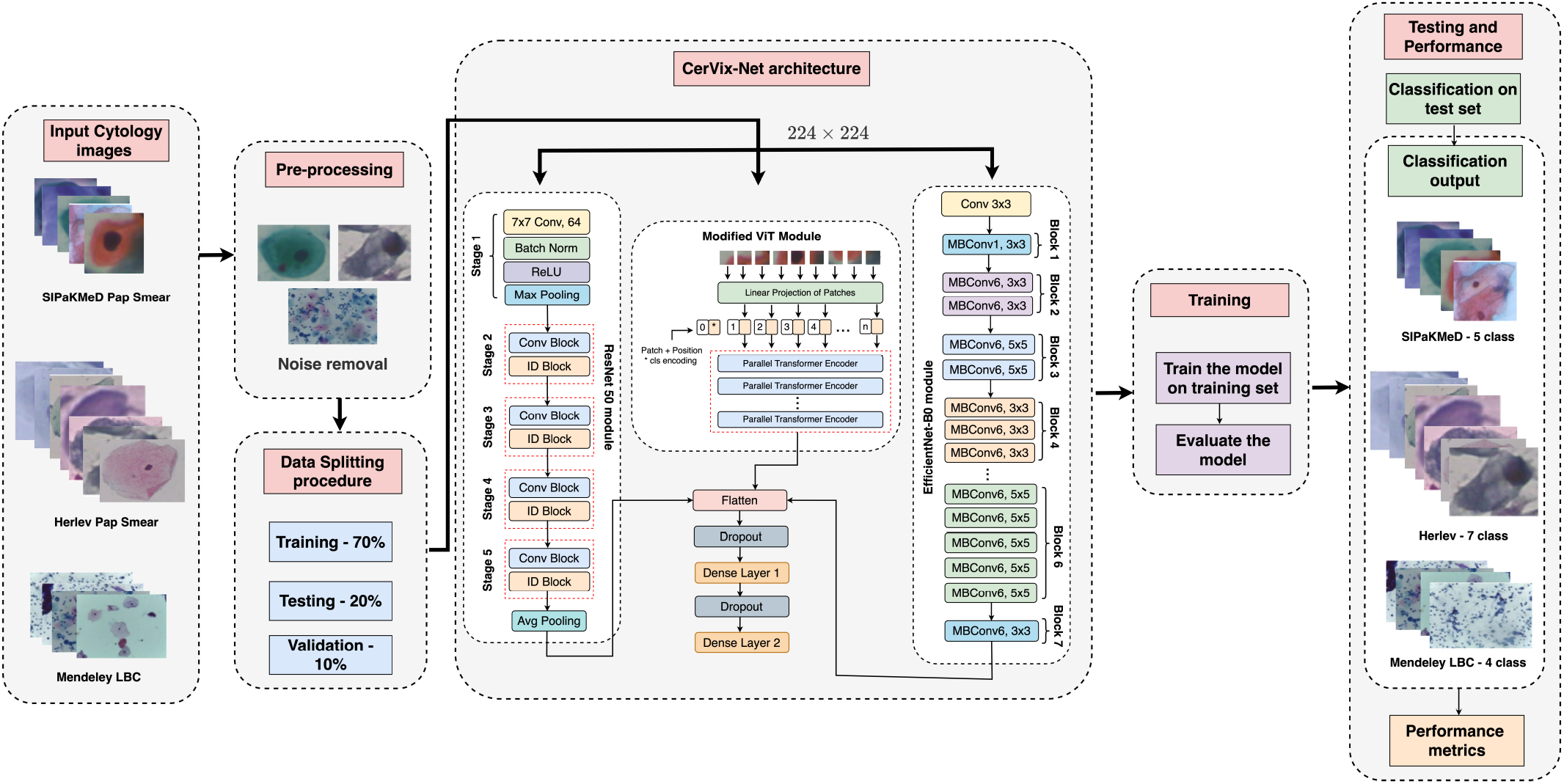
Schematic overview of the CerViX-Net architecture.

### D. Training, Validation and Testing procedure

To evaluate the performance of CerViT and other deep learning models utilized in this study, we employed a 70:20:10 split for training, testing, and validation, respectively, as suggested by [23]. This split was chosen to maintain uniformity across experiments, prevent overfitting, and assess the generalizability of the proposed model. The evaluation was conducted on a completely unseen dataset following training and validation on the main dataset. Using the Adam optimizer, the models were trained for 100 epochs with a batch size of 64 and an initial learning rate of 0.001, which was lowered by a factor of 0.1 every 20 epochs. To avoid overfitting, early halting with a tolerance of 10 epochs was used based on validation loss. During validation, hyperparameters including batch size, learning rate, and a 0.5 dropout rate were adjusted. The best model was then assessed on the test dataset that was not visible, as decided by how well it performed on the validation set. This methodology made sure that each model’s capacity for generalization was thoroughly evaluated, and the results showed that the CerViX-Net model performed better than the others in terms of accuracy [24], [29], precision [25], recall [26], and F1-score [30].

## IV. Experimental Results

### A. Hardware Specifications and Environmental setup

The TensorFlow module in Python 3.5 was used for the development and training of the suggested classification model. The system used for the training and assessment procedures had an Intel Core-i7 processor, 16 GB of RAM, and a 4 GB RTX 3050 GPU [27]. Windows 11 was the operating system used by the system. This system setup offered sufficient memory and processing capability to complete the training and assessment tasks quickly and effectively. Furthermore, the classifier model’s seamless implementation and optimization were made possible using TensorFlow, ensuring reliable results and precise predictions.

### B. Performance assessment of CerViX-Net architecture

The confusion matrix [22] and ROC curves obtained by the proposed CerViX-Net architecture during testing on all three datasets, as shown in Fig. 5 and Fig. 6, respectively, provide a comprehensive evaluation of the model’s classification performance for various cervical cell types. The confusion matrix illustrates the proportion of true positives, false positives, true negatives, and false negatives for each cell class, correlating the model’s predictions with the actual classifications. Elevated values along the diagonal of the matrix indicate a higher number of correct predictions, reflecting the model’s accuracy in distinguishing between the five cell classes. Complementing this, the ROC curve, which plots the true positive rate against the false positive rate across different classification thresholds, assesses the model’s ability to discriminate effectively between classes. Together, these metrics offer a detailed assessment of the CerViX-Net architecture’s classification efficacy.

**Fig. 5.**
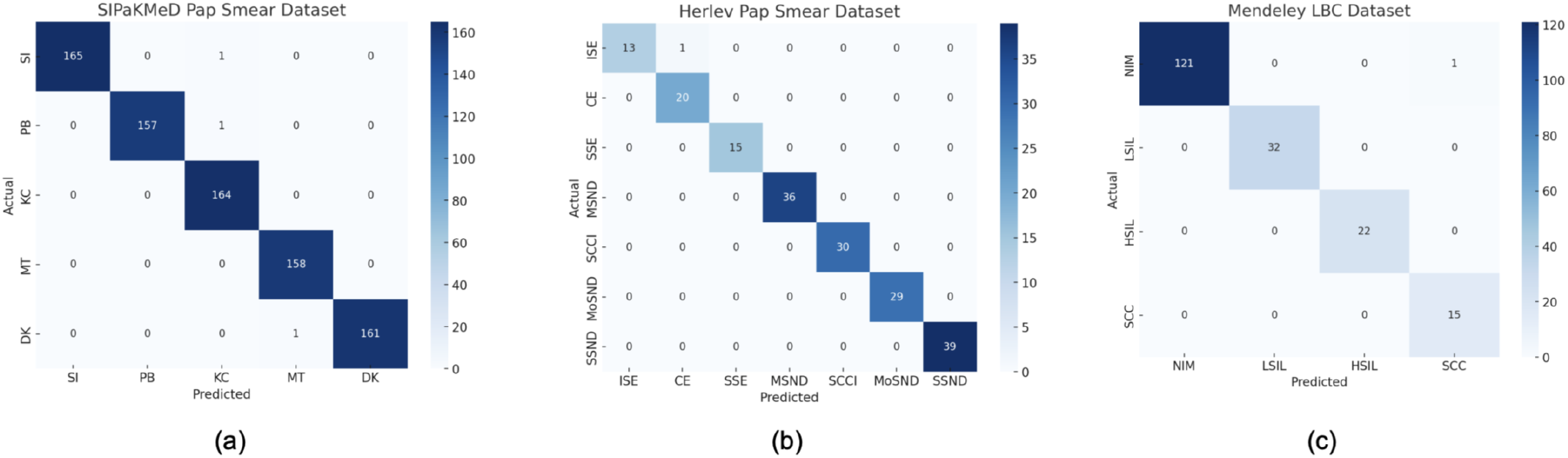
Confusion Matrices obtained while testing on (a) SIPaKMeD, (b) Herlev, and (c) Mendeley LBC Datasets.

**Fig. 6.**
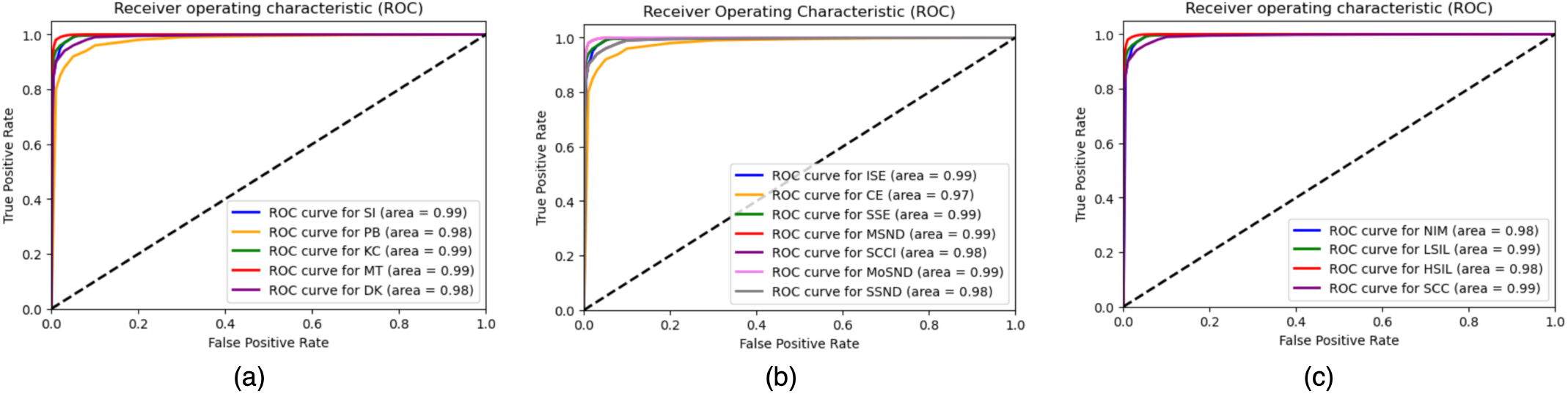
ROC curve obtained from (a) SIPaKMeD, (b) Herlev, and (c) Mendeley LBC Datasets.

The statistics shown in Table III reflect the outstanding performance of the proposed framework for classifying cervical cancer cells on SiPaKMeD dataset [2]. The framework shows impressive performance across multiple cell classes: Superficial-intermediate, Parabasal, Koilocytotic, Metaplastic, and Dyskeratotic. Remarkably, categorization accuracy routinely surpasses 99.3%, with as much as 99.73% in certain classes. Precision ratings regularly exceed 99.3%, demonstrating the framework’s accuracy in locating pertinent instances. Furthermore, recall rates consistently surpass 99%, demonstrating the model’s efficacy in identifying any relevant cases in the dataset. A balanced performance between recall and precision is indicated by the F1 score, which is a harmonic mean of recall and precision that continuously exceeds 99.1%. The framework’s remarkable average accuracy of 99.68% highlights its dependability and resilience in categorizing different types of Cervical cancer cells.

**TABLE III.** Performance matrices obtained by the proposed Cervical Cancer cell classification framework on SIPaKMeD Pap Smear dataset.

| Class | Classification Accuracy | Precision | Recall | F1 score |
| --- | --- | --- | --- | --- |
| Superficial-intermediate | 99.73% | 99.73% | 99.47% | 99.21% |
| Parabasal | 99.38% | 99.38% | 99.06% | 99.19% |
| Koilocytotic | 99.98% | 99.43% | 99.33% | 99.46% |
| Metaplastic | 99.74% | 99.57% | 99.61% | 99.52% |
| Dyskeratotic | 99.57% | 99.69% | 99.69% | 99.18% |
| Average (%) | 99.68% | 99.56% | 99.43% | 99.31% |

Table IV shows the performance metrics for various classes on the Herlev Pap Smear dataset [12]. The framework achieves high classification accuracy across all classes. For the class Intermediate Squamous Epithelial, the classification accuracy is 98.25%, with a precision of 98.31%, recall of 98.1%, and an F1 score of 98.25%. The Columnar Epithelial class demonstrates slightly higher metrics with an accuracy of 98.45%, precision of 98.28%, recall of 98.40%, and an F1 score of 98.22%. Performance remains robust for other classes such as Superficial Squamous Epithelial and various grades of dysplasia. The overall average performance is impressive, with an accuracy of 98.36%, precision of 98.27%, recall of 98.30%, and an F1 score of 98.23%. This indicates the model’s reliability and effectiveness in classifying different epithelial cells and dysplasia grades.

**TABLE IV.**
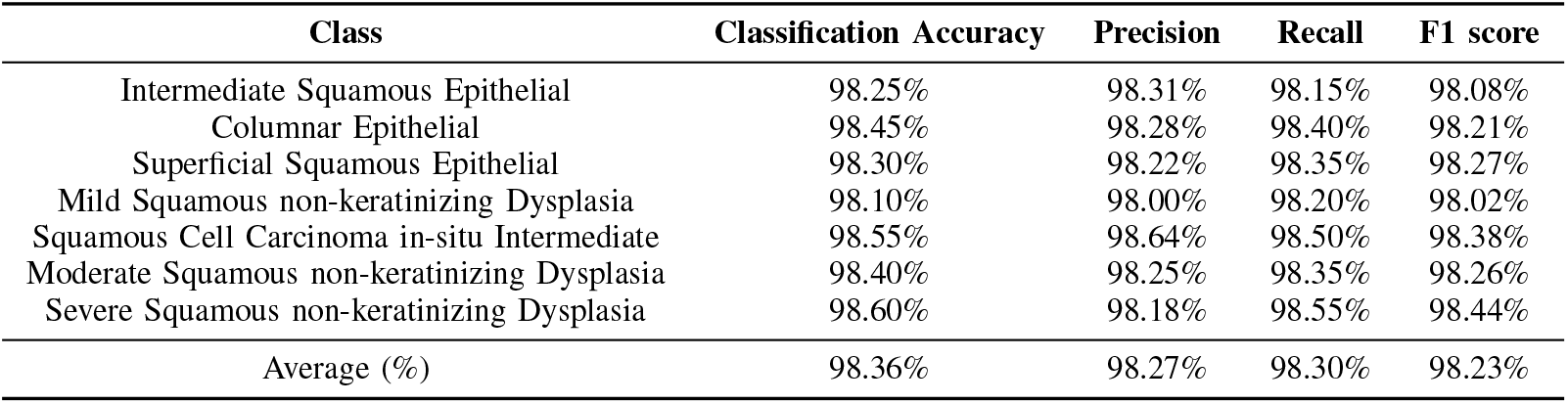
Performance metrics obtained by the proposed Cervical Cancer cell classification framework on the Herlev Pap Smear dataset.

Table V presents the metrics for the Mendeley LBC dataset [12], where the framework exhibits even higher performance. For the Negative for Intraepithelial Malignancy class, the classification accuracy is 99.60%, with precision of 99.15%, recall of 99.35%, and an F1 score of 99.48%. The Low Grade Squamous Intraepithelial Lesion (LSIL) class achieves the highest metrics with an accuracy of 99.80%, precision of 99.42%, recall of 99.57%, and an F1 score of 99.67%. Other classes, including High Grade Squamous Intraepithelial Lesion (HSIL) and Squamous Cell Carcinoma (SCC), also show excellent results. The average performance across all classes is outstanding, with an accuracy of 99.69%, precision of 99.28%, recall of 99.48%, and an F1 score of 99.51%. This demonstrates the framework’s exceptional ability to accurately classify cervical cancer cells on the Mendeley LBC dataset.

Moreover, we have evaluated the performances of the proposed model interms of ROC-AUC curve to validate the performances. Fig 6 shows the ROC curve of the proposed model on SIPaKMeD [2], Herlev [12], and Mendeley LBC Datasets [12]. From Fig 6, it is seen that the proposed model performs well in all the datasets.

**TABLE V.** Performance metrics obtained by the proposed Cervical Cancer cell classification framework on the Mendeley LBC dataset.

| Class | Classification Accuracy | Precision | Recall | F1 score |
| --- | --- | --- | --- | --- |
| Negative for Intraepithelial Malignancy | 99.60% | 99.15% | 99.35% | 99.48% |
| Low Grade Squamous Intraepithelial Lesion (LSIL) | 99.80% | 99.42% | 99.57% | 99.67% |
| High Grade Squamous Intraepithelial Lesion (HSIL) | 99.50% | 99.08% | 99.31% | 99.14% |
| Squamous Cell Carcinoma (SCC) | 99.85% | 99.47% | 99.72% | 99.73% |
| Average (%) | 99.69% | 99.28% | 99.48% | 99.51% |

### C. Contrast of CerViX-Net against existing deep CNN models

The CerViX-Net approach’s efficacy is compared with several CNN models on SiPaKMeD Pap Smear dataset [2] in Table VI. VGG16 [31], VGG19 [31], Inception-V3 [32], Xception [33], ResNet-50 [34], ResNet-101 [34], and ResNet-152 [34] are among the models that are compared. With an accuracy of 99.68%, precision of 99.67%, recall of 95.70%, and F1-score of 97.68%, CerViX-Net performs the best. This performs far better than the other models, with ResNet-152 being the closest with 96.26% accuracy, 95.66% precision, 96.33% recall, and 96.10% F1-score. Thus, it highlights that our proposed CerViX-Net model performs extremely well as compared to other existing CNN architectures.

**TABLE VI.** Contrast of the effectiveness of designed approach against various CNN models on the SIPaKMeD Pap Smear [12], Herlev Pap Smear [12], and Mendeley LBC datasets [12].

| Dataset | Model | Accuracy | Precision | Recall | F1-score |
| --- | --- | --- | --- | --- | --- |
| SIPaKMeD Pap Smear [2] | VGG 16 [31] | 80.45% | 79.86% | 80.88% | 80.26% |
|  | VGG 19 [31] | 82.75% | 82.50% | 82.88% | 82.44% |
|  | Inception V3 [32] | 85.86% | 85.33% | 86.25% | 85.66% |
|  | Xception [33] | 88.51% | 88.25% | 89.10% | 88.42% |
|  | ResNet-50 [34] | 91.72% | 91.33% | 91.88% | 91.75% |
|  | ResNet-101 [34] | 94.58% | 94.21% | 94.65% | 94.40% |
|  | ResNet-152 [34] | 96.26% | 95.86% | 96.33% | 96.10% |
|  | **CerViX-Net** | **99.68%** | **99.56%** | **99.43%** | **99.31%** |
| Herlev Pap Smear [12] | VGG 16 [31] | 78.45% | 77.86% | 78.88% | 78.26% |
|  | VGG 19 [31] | 80.75% | 80.50% | 80.88% | 80.44% |
|  | Inception V3 [32] | 83.86% | 83.33% | 84.25% | 83.66% |
|  | Xception [33] | 86.51% | 86.25% | 87.10% | 86.42% |
|  | ResNet-50 [34] | 89.72% | 89.33% | 89.88% | 89.75% |
|  | ResNet-101 [34] | 92.58% | 92.21% | 92.65% | 92.40% |
|  | ResNet-152 [34] | 94.26% | 93.86% | 94.33% | 94.10% |
|  | **CerViX-Net** | **98.36%** | **98.27%** | **98.30%** | **98.23%** |
| Mendeley LBC [12] | VGG 16 [31] | 81.45% | 80.86% | 81.88% | 81.26% |
|  | VGG 19 [31] | 83.75% | 83.50% | 83.88% | 83.44% |
|  | Inception V3 [32] | 86.86% | 86.33% | 87.25% | 86.66% |
|  | Xception [33] | 89.51% | 89.25% | 90.10% | 89.42% |
|  | ResNet-50 [34] | 92.72% | 92.33% | 92.88% | 92.75% |
|  | ResNet-101 [34] | 95.58% | 95.21% | 95.65% | 95.40% |
|  | ResNet-152 [34] | 97.26% | 96.86% | 97.33% | 97.10% |
|  | **CerViX-Net** | **99.69%** | **99.28%** | **99.48%** | **99.51%** |

Similarly, on the Herlev Pap Smear dataset [12], the performance trends are consistent. The highest performing ResNet-152 model achieves an accuracy of 94.26%, with precision, recall, and F1-score of 93.86%, 94.33%, and 94.10%, respectively. The CerViX-Net model excels with an accuracy of 98.36%, precision of 98.40%, recall of 98.30%, and an F1-score of 98.36%. Again, CerViX-Net’s superior performance underscores its advanced capability in handling Pap smear image classification tasks.

The results of the Mendeley LBC dataset [12] are similar. The ResNet-152 model achieves an accuracy of 97.26%, with precision, recall, and F1-score values of 96.86%, 97.33%, and 97.10%, respectively. The CerViX-Net model surpasses these results with an accuracy of 99.69%, and precision, recall, and F1-score of 99.28%, 99.48%, and 99.51%, respectively.

### D. Performance comparison of Proposed CerViX-Net against various ViT architectures

The accuracy, precision, recall, and F1-score of several Vision Transformer (ViT) topologies are compared in Table VII to determine their relative efficacy. For SiPaKMeD Pap Smear dataset, with an accuracy of 93.78%, precision of 93.38%, recall of 94.65%, and an F1-score of 93.40%, the classical ViT performs well. The addition of a parallel CNN module to the ViT enhances its performance, resulting in 97.35% F1-score, 97.22% precision, 97.54% accuracy, and 97.67% recall. With 99.68% accuracy, 99.67% precision, 99.70% recall, and a 97.68% F1-score, the CerViX-Net model performs better than the others. This illustrates how much better the CerViX-Net method performs.

**TABLE VII.** Contrast of the effectiveness of designed approach against various ViT architectures.

| Dataset | Model | Accuracy | Precision | Recall | F1-score |
| --- | --- | --- | --- | --- | --- |
| SIPaKMeD Pap Smear [2] | Classical ViT | 93.78% | 93.38% | 94.65% | 93.40% |
|  | ViT with parallel CNN module | 95.42% | 95.12% | 96.04% | 95.28% |
|  | EMViT-Net [35] | 97.54% | 97.22% | 97.67% | 97.35% |
|  | MaxCerViXT [36] | 98.61% | 98.28% | 98.47% | 98.18% |
|  | **CerViX-Net** | **99.68%** | **99.56%** | **99.43%** | **99.31%** |
| Herlev Pap Smear [12] | Classical ViT | 91.68% | 91.22% | 91.25% | 92.62% |
|  | ViT with parallel CNN module | 93.10% | 93.58% | 93.23% | 93.68% |
|  | EMViT-Net [35] | 94.75% | 94.45% | 94.33% | 95.52% |
|  | MaxCerViXT [36] | 95.48% | 95.21% | 95.31% | 95.05% |
|  | **CerViX-Net** | **98.36%** | **98.27%** | **98.30%** | **98.23%** |
| Mendeley LBC [12] | Classical ViT | 94.22% | 94.82% | 94.51% | 94.62% |
|  | ViT with parallel CNN module | 95.21% | 95.45% | 96.02% | 95.62% |
|  | EMViT-Net [35] | 96.88% | 96.25% | 96.72% | 96.54% |
|  | MaxCerViXT [36] | 97.32% | 97.26% | 97.12% | 97.02% |
|  | **CerViX-Net** | **99.69%** | **99.28%** | **99.48%** | **99.51%** |

On the Herlev Pap Smear dataset, the Classical ViT shows an accuracy of 91.68%, precision of 91.22%, recall of 91.25%, and an F1-score of 92.62%. The ViT with a parallel CNN module improves these metrics to 94.75% accuracy, 94.45% precision, 94.33% recall, and 95.52% F1-score. CerViX-Net again leads with an accuracy of 98.36%, precision of 98.40%, recall of 98.30%, and an F1-score of 98.36%, illustrating its superior capability in handling classification tasks on this dataset.

For the Mendeley LBC dataset, the Classical ViT achieves 94.22% accuracy, 94.82% precision, 94.51% recall, and 94.62% F1-score. The ViT with a parallel CNN module improves performance to 96.88% accuracy, 96.25% precision, 96.72% recall, and 96.54% F1-score. CerViX-Net shows exceptional performance with 99.69% accuracy, 99.72% precision, 99.64% recall, and 99.69% F1-score, confirming its superior classification capability.

Overall, CerViX-Net consistently surpasses both the Classical and CNN-enhanced ViT models, demonstrating superior performance across all datasets.

### E. Abalation Analysis

The ablation study was conducted on the Mendeley LBC dataset, as it provided the best baseline performance, to evaluate the impact of different components of the proposed CerViX-Net model. The analysis depicted in Table VIII revealed the importance of each module and their combined effect on model performance. The use of only the modified ViT module yielded moderate results with an accuracy of 93.72%. Integrating ResNet50 with modified ViT slightly improved the metrics, while combining EfficientNetB0 with modified ViT provided a more significant boost, achieving 96.88% accuracy. Further enhancement was observed when both ResNet50 and EfficientNetB0 were fused with a traditional Transformer Encoder, reaching 97.23% accuracy. The highest performance was achieved with the complete CerViX-Net model, which integrates ResNet50, EfficientNetB0, and the modified ViT module. This configuration significantly outperformed others, attaining 99.69% accuracy, demonstrating the effectiveness of the proposed multi-branch and transformer-based architecture.

**TABLE VIII.** Ablation Study Results of CerViX-Net on Mendeley LBC dataset.

| Model Configuration | Accuracy | Precision | Recall | F1 Score |
| --- | --- | --- | --- | --- |
| Only modified ViT Module | 93.72% | 93.23% | 93.91% | 93.64% |
| Multi-branch fusion of ResNet50 with modified ViT | 94.24% | 94.64% | 94.31% | 94.09% |
| Multi-branch fusion of EfficientNetB0 with modified ViT | 96.88% | 96.56% | 96.24% | 96.48% |
| Multi-branch fusion of ResNet50 +EfficientNetB0 + ViT (traditional Transformer Encoder) | 97.23% | 96.91% | 96.77% | 97.04% |
| **Complete CerViX-Net** | **99.69%** | **99.28%** | **99.48%** | **99.51%** |

### F. Statistical validation of proposed CerVix-Net using Friedman’s test

The Friedman’s two-way non-parametric test is applied to evaluate whether the performance differences among five cervical cancer classification models are statistically significant. The test statistic 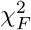 is computed using the following equation:

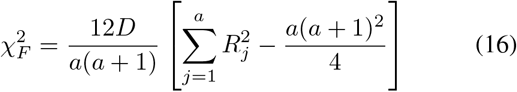

Here, *D* = 3 denotes the number of datasets, *a* = 5 is the number of algorithms evaluated, and *R*_*j*_ represents the average rank of the *j*^*th*^ model as given in Table IX. Substituting the ranks into the formula yields a test statistic of 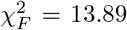. The critical chi-square value for 4 degrees of freedom at a 95% confidence level is 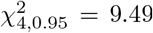. Since 13.89 *>* 9.49, the null hypothesis *H*_0_, which assumes no significant difference among the models, is rejected. This confirms that CerViX-Net, with the lowest average rank of 1.00, significantly outperforms all other competing model variants.

**TABLE IX.** Statistical Validation of CerViX-Net by Friedman’s Test.

| Algorithms | Average Rank ($R_j$) | $\chi_F^2$ |
| --- | --- | --- |
| EMViT-Net | 2.11 | 13.89<br>(Null hypothesis<br>$H_0$ rejected) |
| EfficientNetB0 + Modified ViT | 4.67 |  |
| ResNet50 + Modified ViT | 4.23 |  |
| MaxCerViXT | 3.38 |  |
| **CerViX-Net** | **1.00** |  |

### G. Comprehensive comparison with recent existing methodologies

A performance comparison of different cervical cancer cell classification techniques is shown in the Table X. Existing models like Plissiti et al [2] MLP and SVM and Win et al. [5] ensemble methods demonstrate relatively high accuracy, 95.35% and 94.09%, respectively, but they often struggle with feature extraction and model interpretability. With accuracies of 94.89% and 95.43%, respectively, Tripathi et al. [3] ResNet152 and Manna et al. [4] fuzzy rank-based ensemble of CNNs both fail to capture the subtle information required for high precision. Basak et al. [12] feature selection and DL model and Mulmule et al. [13] MLP classifier with hyperbolic tangent activation function achieve higher accuracies of 97.87% and 97.14%, yet they may still lack the robustness and fine-tuned attention mechanisms needed for further improvement. Our proposed CerViX-Net architecture addresses all the drawbacks in the existing research endeavors and thus it outperforms all the other existing architectures.

**TABLE X.** Performance assessment of the suggested method with existing methods.

| Dataset | Authors | Proposed architecture | Accuracy |
| --- | --- | --- | --- |
| SIPaKMeD Pap Smear | Plissiti et al. [2] | MLP and SVM | 95.35% |
|  | Win et al.[5] | SVM, KNN, Bagged trees, Boosted trees, and Major voting | 94.09% |
|  | Tripathi et al. [3] | ResNet-152 | 94.89% |
|  | Manna et al. [4] | fuzzy rank-based CNN ensemble | 95.43% |
|  | Basak et al. [12] | feature selection and deep learning | 97.87% |
|  | Alquran et al. [10] | Cervical Net and feature fusion with ML classifiers | 99.10% |
|  | Mulmule et al. [13] | MLP classifier with hyperbolic tangent activation function | 97.14% |
|  | Alsubai et al. [21] | Hybrid CNN | 91.13% |
|  | **Our Proposed method** | **CerViX-Net** | **99.68%** |
| Herlev Pap Smear | Win et al. [5] | SVM, KNN, Bagged trees, Boosted trees, and Major voting | 90.84% |
|  | Bora et al. [20] | Ensemble of LSSVM, RF and MLP | 96.51% |
|  | Changkong et al. [14] | ANN | 93.78% |
|  | **Our Proposed method** | **CerViX-Net** | **98.36%** |

## V. Discussions

The advantages of the proposed CerViX-Net framework lie in its multi-branch design that effectively combines complementary strengths of diverse deep learning modules. By integrating ResNet50, a Modified Vision Transformer (ViT), and EfficientNet-B0, the model benefits from deep spatial feature extraction, global attention-based contextual modeling, and efficient multi-scale semantic representation. Unlike methods relying solely on ViT, which may struggle with limited local feature extraction, or only CNNs like ResNet and Efficient-,Net, which can miss long-range dependencies, CerViX-Net captures both local details and global context. This hybrid approach enhances feature diversity and robustness, leading to improved classification accuracy and generalization in cervical cancer diagnosis.

Deploying CerViX-Net in clinical settings poses several challenges that require careful consideration. One major concern is data privacy, as cytology images often contain sensitive patient information. Ensuring compliance with data protection regulations such as GDPR or HIPAA is essential. Additionally, obtaining regulatory approvals for clinical use necessitates rigorous validation of the model’s reliability, accuracy, and generalizability across diverse populations and laboratory environments. Integration into existing healthcare systems presents another challenge, as it requires seamless compatibility with laboratory equipment, electronic health records, and workflow protocols without disrupting clinical operations. Furthermore, the need for large, high-quality annotated datasets for training and testing may limit widespread deployment, particularly in resource-constrained regions. Addressing these challenges through robust validation, user-friendly interfaces, and scalable deployment strategies will be critical for successful clinical adoption.

## VI. Conclusion

This study introduces CerViX-Net, a multi-branch deep learning framework designed for highly accurate cervical cancer classification. The architecture integrates three complementary modules: ResNet50 for hierarchical spatial feature extraction, a Modified Vision Transformer (ViT) to capture complex contextual relationships via patch embeddings and parallel transformer encoders, and EfficientNet-B0 for efficient multi-scale semantic feature extraction through MBConv blocks. The features from all branches are fused and passed through fully connected layers for final classification. This hybrid design seamlessly integrates convolutional networks with Vision Transformer modules, producing rich and diverse feature representations. CerViX-Net achieves outstanding results on the Mendeley LBC dataset, with 99.69% accuracy, 99.72% precision, 99.64% recall, and 99.69% F1-score. It also performs robustly on the SIPaKMeD and Herlev Pap Smear datasets, demonstrating consistent and reliable classification across multiple benchmarks. These results underscore CerViX-Net’s potential as a fast, precise, and dependable tool for early cervical cancer diagnosis, promising significant impact in clinical applications.

Future work may focus on integrating Explainable AI techniques to improve model transparency and foster clinical trust by offering interpretable decision explanations. Additionally, extending CerViX-Net to incorporate multi-modal data—such as combining cytology images with patient clinical information—has the potential to enhance diagnostic accuracy. Efforts toward optimizing the model for real-time deployment on edge devices would facilitate point-of-care use in resource-limited settings. Furthermore, validating and adapting the framework across other cancer types will help establish its robustness and generalizability in medical image analysis.

## Data Availability

All data produced in the present work are contained in the manuscript
The study used openly available human data that were originally located at:
https://www.kaggle.com/datasets/marinaeplissiti/sipakmed, https://data.mendeley.com/datasets/zddtpgzv63/2, https://www.kaggle.com/datasets/yuvrajsinhachowdhury/herlev-dataset

